# Effect of tenofovir on the outcomes of COVID-19 in persons with chronic hepatitis B: a nationwide cohort study in Sweden

**DOI:** 10.64898/2026.06.10.26355365

**Authors:** Frida Jakobsson, Marie Eriksson, Sebastian Kalucza, Anne-Marie Fors Connolly

## Abstract

**Background:** Patients with chronic hepatitis B (CHB) may have an increased risk of severe COVID-19. Tenofovir has been hypothesized to confer protection against severe disease, but evidence is inconclusive. We evaluated the risk of severe COVID-19 among CHB patients treated with tenofovir compared with other nucleos(t)ide analogues (NAs).

**Methods and findings:** In this nationwide, registry-based cohort study, we included all adults with CHB and laboratory-confirmed COVID-19 in Sweden between February 2020 and July 2022. Data from national health and socioeconomic registers were linked using unique personal identification numbers (PINs). Patients with HIV, hepatitis C, or hepatitis D coinfection were excluded. Exposure was defined as tenofovir versus other NA therapy. The primary outcome was severe COVID-19, defined as hospitalization >2 days or death within 30 days of diagnosis. Logistic regression was used to estimate adjusted odds ratios (aOR) with 95% confidence intervals (CI), controlling for age, sex, comorbidities, vaccination, socioeconomic status, and region of birth.

Among 5,877 CHB patients with COVID-19, 672 were receiving NA therapy (437 tenofovir, 235 other NAs). Severe COVID-19 occurred in 8.0% of tenofovir-treated patients and 14.5% of those receiving other NAs (unadjusted OR 0.52; 95% CI, 0.31-0.85). After adjustment, the association was attenuated and no longer significant (aOR 0.72; 95% CI, 0.39–1.31). Older age, comorbidities, and unvaccinated status were strongly associated with severe disease.

**Conclusions:** The apparent protective effect of tenofovir against severe COVID-19 in unadjusted analyses was largely explained by confounding factors. The risk of severe disease was primarily driven by age, comorbidities, and vaccination status. Prevention of severe COVID-19 in patients with CHB should instead focus on vaccination and management of comorbidities.

## Introduction

As of January 2026, the Coronavirus disease 2019 (COVID-19) pandemic has caused more than 7,1 million deaths worldwide, according to the World Health Organization (WHO) (1). Although widespread vaccination and the introduction of antiviral therapies have brought the global epidemic largely under control, individuals with underlying conditions remain at elevated risk for severe disease (2). Evidence suggests that patients with CHB may be vulnerable to severe COVID-19 (3–5). However, whether antiviral treatment for Hepatitis B virus (HBV) infection confers any protective effect against severe COVID-19 remains uncertain. Numerous pharmacologic agents have been investigated for prophylaxis and treatment of COVID-19, but most have shown limited or inconclusive evidence of clinical benefit(6–9).

Globally, an estimated 240 million people live with CHB, and many are eligible for therapy. Currently, there are two main treatment options for CHB patients: treatment with a NA or with Interferon-alpha (IFNa)(10). The NAs that have been approved in Europe for HBV treatment include lamivudine (LAM), adefovir dipivoxil (ADV), entecavir (ETV), telbivudine (TBV), tenofovir disoproxil fumarate (TDF) and tenofovir alafenamide (TAF)(10). The most frequently used HBV treatments are ETV, TDF and TAF because of their long-term effectiveness and tolerability(10, 11). ETV may be less effective in patients with LAM-resistant HBV-strains(12, 13), and TDF is associated with impaired renal function and reductions in bone mineral density(14). TAF, a relatively new tenofovir prodrug, has been developed to overcome the less favourable safety profile of TDF(15). In Sweden tenofovir (TDF/TAF) and ETV are the first line treatment options for hepatitis B patients(16). Recently, several studies reported that, tenofovir, as the first-line anti- HBV drug, could reduce the risk of severe COVID-19 (17–21) but there is controversy about these findings. To date, most studies have been limited by small sample sizes and a lack of data on population diversity, including ethnicity and socioeconomic status, which may influence both the risk for severe COVID-19 and treatment outcomes(19, 21–24) Most of these studies include patients with HIV, or healthy populations, but not many have focused specifically on HBV-infected individuals(8, 25). Clarifying whether tenofovir provides additional protection against severe COVID-19 could have important implications for HBV- treatment decisions in this population.

Therefore, the objective of this study was to evaluate the severity of COVID-19 in patients with CHB receiving antiviral therapy, with the aim of determining whether tenofovir conferred protective effects against severe COVID-19 compared to other NAs.

## Methods

### Source of data and study population

This register-based nationwide study was conducted in Sweden with data from 1 February 2020 until 27 July 2022. The study population included all adult CHB patients in Sweden, treated with NA, at COVID-19 diagnosis. COVID-19 and Hepatitis B are both notifiable diseases in Sweden, and all diagnosed individuals are reported to SmiNet (Swedish Public Health Agency). Data for hepatitis B diagnosis, COVID-19 diagnosis and COVID-19 vaccination were retrieved from Public Health Agency of Sweden, where all personal identification numbers (PINs) were extracted (including all individuals registered as having COVID-19 from Feb 1 2020, until 26 June 2022). The PINs were cross-linked with the following registers administered by the Swedish National Board of Health and Welfare: inpatient register (1987-2022); outpatient register (1997-2022); cancer registry (2000-2022) and cause of death register (2020-2022). Socioeconomic data were obtained by crosslinking the COVID-19 cohort with the Longitudinal Integrated Database for Health Insurance and Labor Market Studies (LISA) registry, from Statistics Sweden, using the PINs for each patient. All data were pseudonymised by Statistics Sweden and the National Board of Health and Welfare. To identify the closest date of infection with SARS-CoV-2 from the registry database, we used the earliest of following reported dates: date of disease onset, sample date, diagnosis date, and date of report to SmiNet. A new infection with SARS-CoV 2 was defined as a new infection if it happened >6 months from the previous one.

Chronic HBV-infection is defined as persistence of Hepatitis B surface antigen (HBsAg) in serum for at least 6 months after acute infection. Only patients >17 years old at COVID-19 diagnosis with CHB were included in this study. We excluded patients diagnosed with HIV using ICD codes from in- and outpatient registry (ICD10: B20x, B21x, B22x, B23x, B24x.

ICD9: 042, 043, 044, v08). Hepatitis C virus (HCV) and Hepatitis D virus (HDV) coinfection were also excluded, through diagnosis from SmiNet-register.

### Exposure

The exposure was treatment with TAF/TDF compared to other NA-treatment.

From the prescribed drug registry from Swedish National Board of Health and Welfare all hepatitis B patients treated with NAs were identified using following ATC codes: J05AF05 Lamivudin, J05AF07 Tenofovirdisoproxil, J05AF10 Entekavir, J05AF13 Tenofoviralafenamid, J05AF08 Adefovirdipivoxil, J05AF11 Telbivudin. Only individuals that had an expedited NA 12 months or less before COVID-19 date were included in this study. We excluded individuals with interferon treatment expedited 12 months or less before COVID-19 diagnosis (ATC code L03AB11) (figure 1).

**Figure 1.**
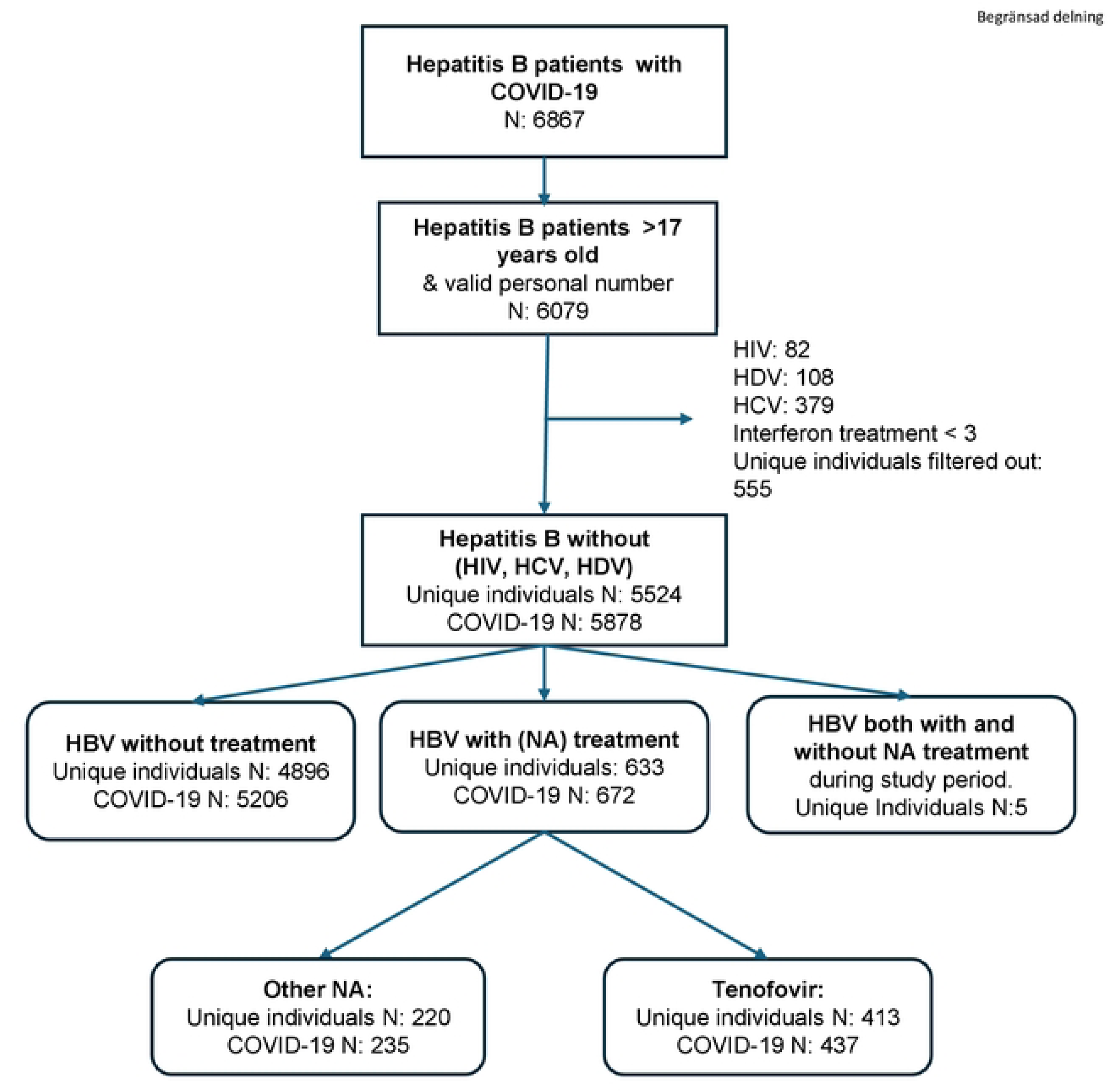
Flowchart of individuals included. Flowchart of number of unique individuals and SARS-CoV2 infections included and excluded in the study. N, number; HDV, hepatitis delta virus; HCV, hepatitis C virus; NA, nucleos(t)ide analogue

### Outcome

The primary outcome of severe COVID-19 was based on a composite measure including hospitalization >2 days or death by COVID-19. Hospitalization or death due to COVID-19 was defined through the ICD10 codes U071 or U072 as main or contributing diagnoses within one month post index date in the inpatient- or cause of death registries, respectively.

### Variables

Covariates included sex, age (at index date), SARS-CoV-2 vaccination status (at least two weeks before index date), co-morbidities, geographic region of birth and socioeconomic factors (income and education level). The weighted Charlson Comorbidity Index (wCCI) was calculated to determine the burden of comorbidities, based on historical registry data from the outpatient register, inpatient register, and the cancer register using a method specifically adapted to Swedish National Registers (26, 27). To ensure complications due to COVID-19 were not included in the wCCI, the calculation for wCCI was stopped one month prior to the index date. If there were no wCCI diagnosis codes in the outpatient register, inpatient register, or the cancer register, the individual was given a wCCI of zero. The country of birth was defined as the birth country registered for that specific individual in the Total Population Registry at Statistics Sweden. The birth countries were divided in two regions, according to international standards based on ISO 3166. The regions were Northern Europe as one group, and rest of the world as the other group (S1 Table). The socioeconomic variables: income and education, were derived from the LISA registry. Income was defined as the annual household income divided into tertiles: lowest, middle, highest. Education was also divided in three stages: primary, secondary, and tertiary education.

### Statistical analysis

Univariable and multivariable logistic regression was used to determine whether tenofovir treatment was associated with a lower risk of severe COVID-19 compared to other NAs. The multivariable analyses included the potential confounders: age (years), sex, region of birth, COVID-19 vaccination (yes/no), comorbidity burden measured by wCCI, education and income. Outcome was presented by OR with 95% confidence intervals (CI). Missing data were handled by adding these as a separate category. Statistical analyses were carried out in R (R Core Team, 2024) using the version 4.4.2. The study received ethical approval from the Swedish Ethical Review Authority (DNR 2020-02150).

## Results

During the study period (1 February 2020 to 27 July 2022), data were available for 6,867 patients with CHB and COVID-19 (Figure 1). After excluding children (<18 years), individuals with invalid personal identification number, individuals co-infected with HIV, HDV, or HCV and those receiving interferon therapy, 5,524 patients remained. During the study period 5,878 SARS-Cov2 infections occurred in the study population.

### Baseline characteristics

Of individuals with COVID-19, 633 received NA-therapy (Figure 1). Those with NA-treatment were older, had more comorbidities, higher burden of cirrhosis (12.6% vs 0.8%) and lower COVID-19 vaccination coverage (37.9% vs 46.1%) compared to hepatitis B individuals without NAs. Both groups had similar socioeconomic status but there were more males with NA-treatment (61.2 vs 49.5%) (Table 1).

**Table 1.** Baseline characteristics of all hepatitis B and SARS-CoV2 infections. Baseline characteristics stratified by ***any nucleoside analog (NA)***. Frequency (%)

|  | No NA<br>(N=5206) | Any NA<br>(N=672) | Overall<br>(N=5878) |
| --- | --- | --- | --- |
| **Severe COVID-19** |  |  |  |
| Non-severe | 4918 (94.5%) | 603 (89.7%) | 5521 (93.9%) |
| Severe | 288 (5.5%) | 69 (10.3%) | 357 (6.1%) |
| **Sex** |  |  |  |
| Male | 2578 (49.5%) | 411 (61.2%) | 2989 (50.9%) |
| Female | 2628 (50.5%) | 261 (38.8%) | 2889 (49.1%) |
| **Age group** |  |  |  |
| 18-49 | 3563 (68.4%) | 393 (58.5%) | 3956 (67.3%) |
| 50-64 | 1341 (25.8%) | 215 (32.0%) | 1556 (26.5%) |
| 65-79 | 274 (5.3%) | 56 (8.3%) | 330 (5.6%) |
| 80+ | 28 (0.5%) | 8 (1.2%) | 36 (0.6%) |
| **Education** |  |  |  |
| Higher Education | 2253 (43.3%) | 285 (42.4%) | 2538 (43.2%) |
| Missing or unknown | 277 (5.3%) | 55 (8.2%) | 332 (5.6%) |
| Primary Education | 1295 (24.9%) | 165 (24.6%) | 1460 (24.8%) |
| Secondary Education | 1381 (26.5%) | 167 (24.9%) | 1548 (26.3%) |
| **Disposable Income** |  |  |  |
| Lowest | 2619 (50.3%) | 339 (50.4%) | 2958 (50.3%) |
| Middle | 1726 (33.2%) | 214 (31.8%) | 1940 (33.0%) |
| Highest | 835 (16.0%) | 118 (17.6%) | 953 (16.2%) |
| Missing or unknown | 26 (0.5%) | 1 (0.1%) | 27 (0.5%) |
| **wCCI_cat** |  |  |  |
| 0-2 | 4507 (86.6%) | 474 (70.5%) | 4981 (84.7%) |
| 3-4 | 230 (4.4%) | 95 (14.1%) | 325 (5.5%) |
| 5+ | 469 (9.0%) | 103 (15.3%) | 572 (9.7%) |
| **Cirrhosis** |  |  |  |
| No | 5163 (99.2%) | 587 (87.4%) | 5750 (97.8%) |
| Yes | 43 (0.8%) | 85 (12.6%) | 128 (2.2%) |
| **COVID-19 vaccination** |  |  |  |
| No | 3234 (62.1%) | 362 (53.9%) | 3596 (61.2%) |
| Yes | 1972 (37.9%) | 310 (46.1%) | 2282 (38.8%) |
| **Region of birth** |  |  |  |
| Northern Europe | 643 (12.4%) | 47 (7.0%) | 690 (11.7%) |
| Rest of the world | 4563 (87.6%) | 625 (93.0%) | 5188 (88.3%) |

NA, nucleos(t)ide analogue; wCCI, weighted Charlson Comorbidity Index; N, number;

Of those treated with NAs, 437 (65%) received tenofovir, while 235 (35%) received other NA agents (Table 2). Of those receiving other NAs, 217 (92.3%) were treated with entecavir (ETV) and 18 (7.7%) with lamivudine (LAM). In both tenofovir-treated and other-NA groups, men predominated (61.3% and 60.9%, respectively). Educational level and income distribution were comparable between groups (Table 2). Tenofovir-treated patients were younger with 33.9% aged ≥50 years vs 52.3% in the other-NA group; and had fewer comorbidities compared with those on other NAs (Table 2). COVID-19 vaccination coverage was slightly lower among tenofovir-treated individuals (43.9% vs 50.2%). These differences were statistically significant for age and CCI. The majority in both groups were born outside Sweden or Northern Europe (Table 2).

**Table 2.** Baseline characteristics of hepatitis B and SARS-Cov2 infections with NA-treatment. Baseline characteristics, stratified by **Tenofovir or other NA** treatment. Frequency (%)

|  | **Other NA**<br>**(N=235)** | **Tenofovir**<br>**(N=437)** | **Overall**<br>**(N=672)** |
| --- | --- | --- | --- |
| **Severe COVID-19** |  |  |  |
| Non-severe | 201 (85.5%) | 402 (92.0%) | 603 (89.7%) |
| Severe | 34 (14.5%) | 35 (8.0%) | 69 (10.3%) |
| **Sex** |  |  |  |
| Male | 143 (60.9%) | 268 (61.3%) | 411 (61.2%) |
| Female | 92 (39.1%) | 169 (38.7%) | 261 (38.8%) |
| **Age group** |  |  |  |
| 18-49 | 106 (45.1%) | 287 (65.7%) | 393 (58.5%) |
| 50-64 | 98 (41.7%) | 117 (26.8%) | 215 (32.0%) |
| 65-79 | 25 (10.6%) | 31 (7.1%) | 56 (8.3%) |
| 80+ | 6 (2.6%) | 2 (0.5%) | 8 (1.2%) |
| **Education** |  |  |  |
| Primary Education | 60 (25.5%) | 114 (26.1%) | 174 (25.9%) |
| Secondary Education | 70 (29.8%) | 135 (30.9%) | 205 (30.5%) |
| Higher Education | 87 (37.0%) | 167 (38.2%) | 254 (37.8%) |
| Missing or unknown | 18 (7.7%) | 21 (4.8%) | 39 (5.8%) |
| **Disposable income** |  |  |  |
| Lowest | 115 (48.9%) | 224 (51.3%) | 339 (50.4%) |
| Middle | 79 (33.6%) | 135 (30.9%) | 214 (31.8%) |
| Highest | 40 (17.0%) | 78 (17.8%) | 118 (17.6%) |
| Missing or unknown | 1 (0.4%) | 0 (0%) | 1 (0.1%) |
| **wCCI_cat** |  |  |  |
| 0-2 | 142 (60.4%) | 332 (76.0%) | 474 (70.5%) |
| 3-4 | 43 (18.3%) | 25 (11.9%) | 95 (14.1%) |
| 5+ | 50 (21.3%) | 53 (12.1%) | 103 (15.3%) |
| **COVID-19 vaccination** |  |  |  |
| No | 117 (49.8%) | 245 (56.1%) | 362 (53.9%) |
| Yes | 118 (50.2%) | 192 (43.9%) | 310 (46.1%) |
| **Region of birth** |  |  |  |
| Northern Europe | 22 (9.4%) | 25 (5.7%) | 47 (7.0%) |
| Rest of world | 213 (90.6%) | 412 (94.3%) | 625 (93.0%) |

NA, nucleos(t)ide analogue; wCCI, weighted Charlson Comorbidity Index; N, number;

### Association between NA and severe COVID-19

In the univariable analysis, tenofovir treatment associated with a reduced risk of severe COVID-19 (OR 0.520, 95% CI 0.310–0.850; *p*=0.009) (Table 3). Age, male sex, co-morbidities, lower socioeconomic status were associated with higher risk for severe COVID-19. Whereas COVID-19 vaccination and birth country outside of northern Europe seemed to have a protective effect in the univariable analysis (Table 3). However, in the multivariable analysis (Table 3, Model 1 and 2; Figure 2), the association between Tenofovir treatment and severe Covid-19 was attenuated and no longer statistically significant (OR: 0.837, 95% CI: 0.479-1.477) after adjustment for differences in sex, age, and CCI, and (OR: 0.716, 95% CI: 0.393-1.313) after further adjustment for vaccination, education, income and region of birth.

**Fig 2.**
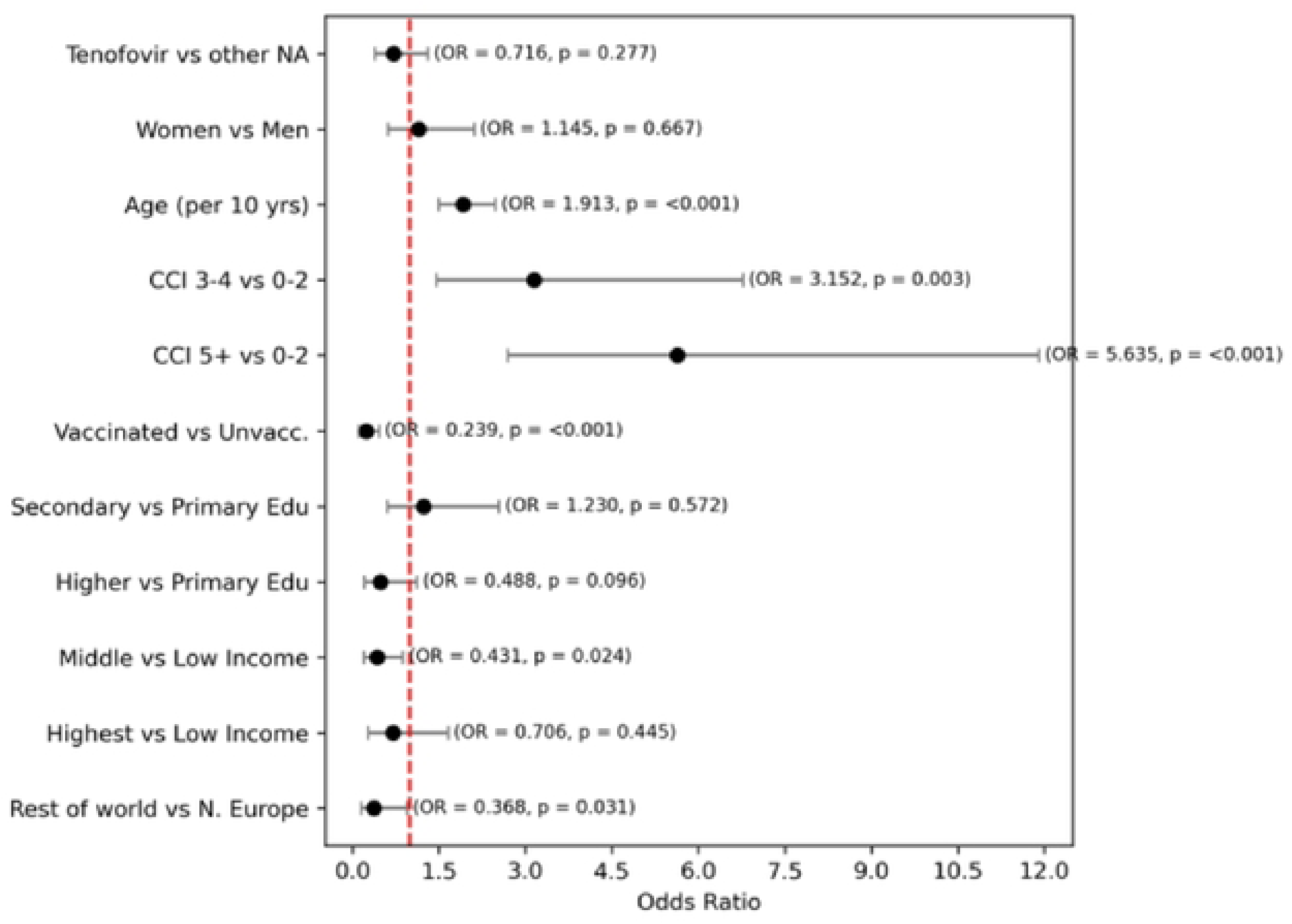
Forest plot. Severe COVID-19 for NA-treated individuals. Logistic regression rnodelling the odds of severe COVID-19 in individuals with N-analog treat,nent. The association, in this rnultivariable model, between Tenofovir treatrnent and severe Covid-19 no longer statistically significant OR: 0.716, 95% CI: 0.393-1.313 after adjustrnent for sex, age, CCI, vaccination, education, incorne and region of birth. OR, odds ratio; NA, nucleos(t)ide analogue; yrs, years; CCI, Charlson Cornorbidity Index; N Europe, Northern Europe; vs, versus; Edu, education.

**Table 3.**
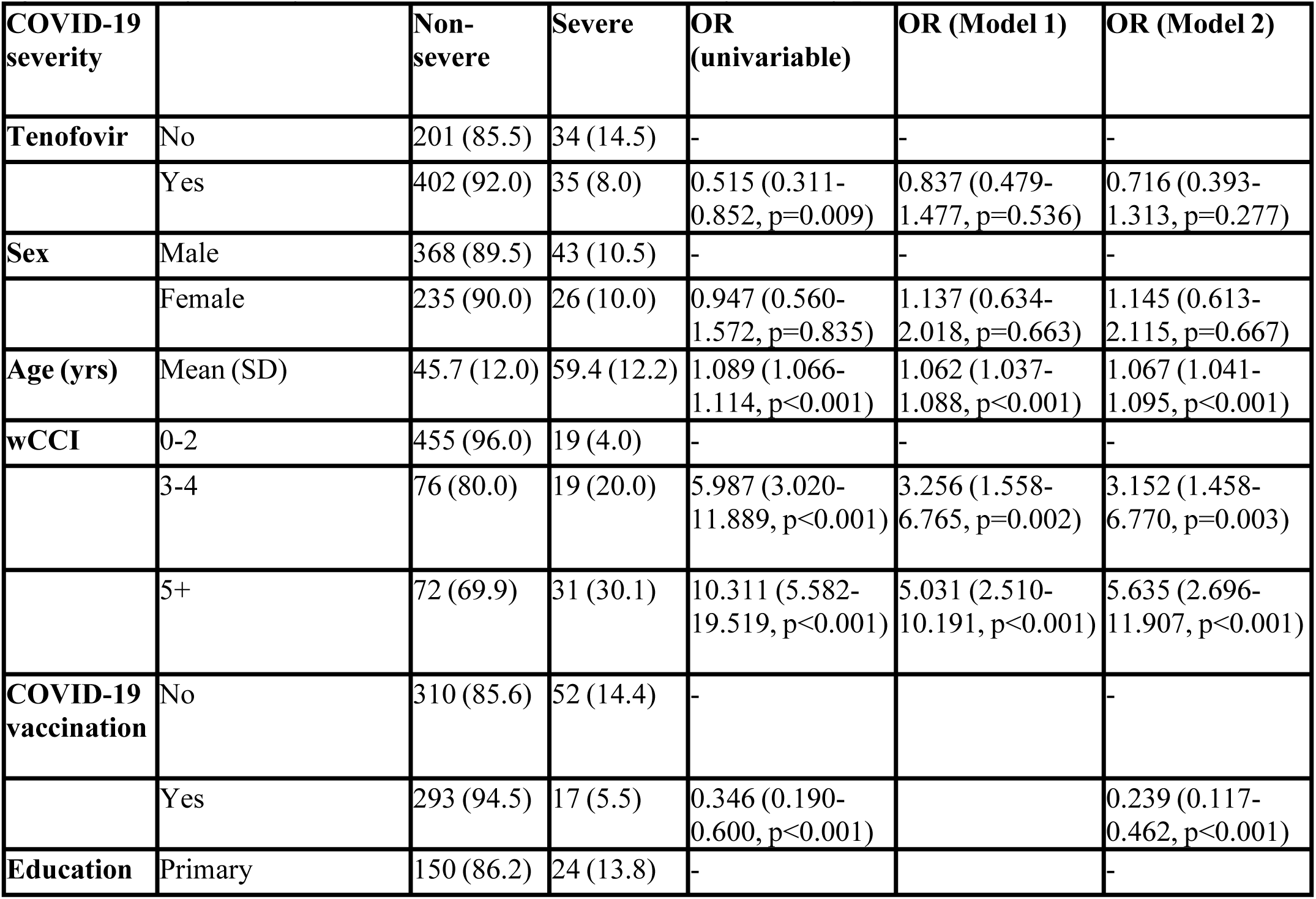
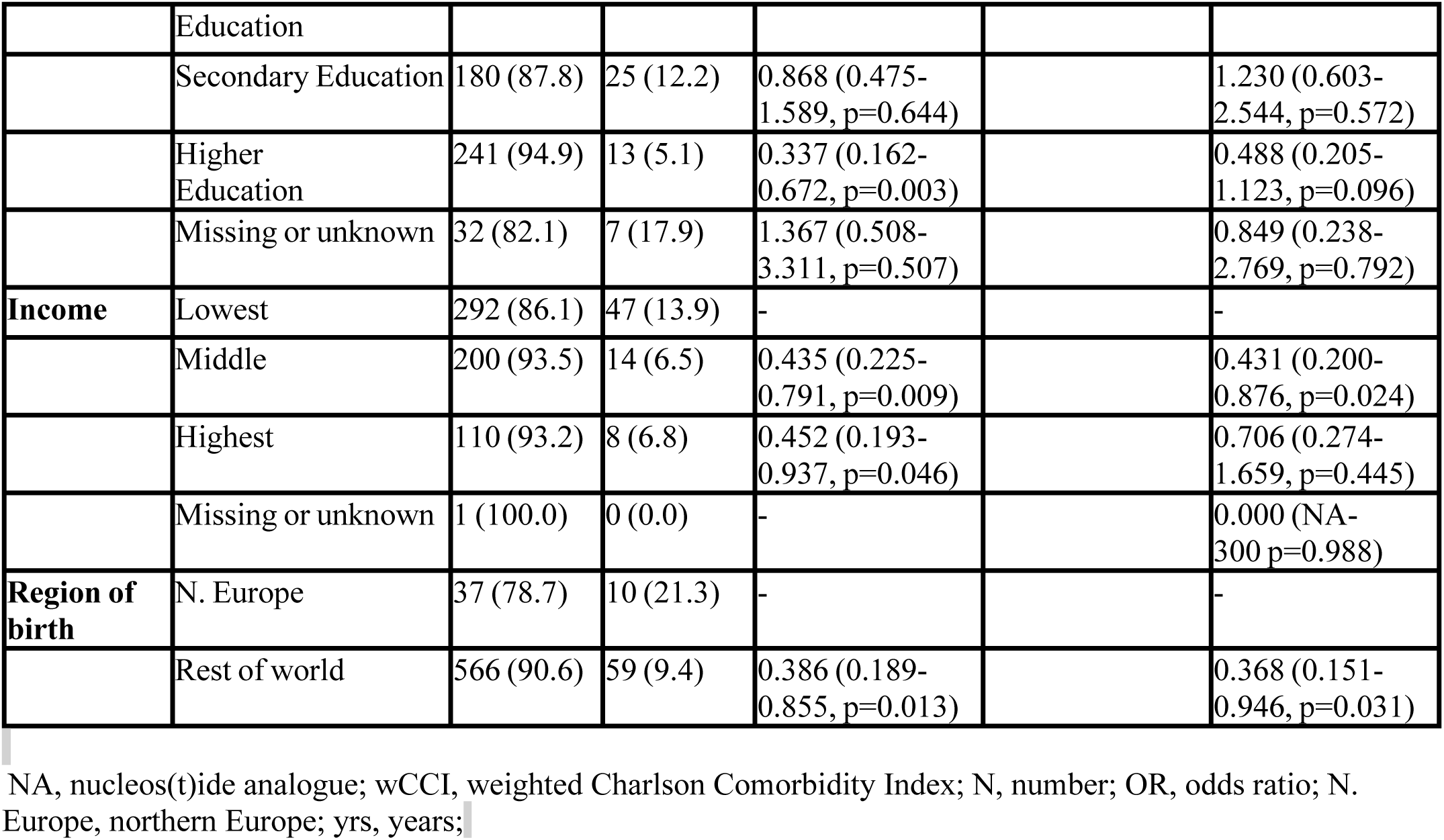
Severe COVID-19 for NA-treated individuals. Logistic regression modelling the odds of severe COVID-19 in individuals with N-analog treatment. Separate univariable models for each independent variable, and multivariable models adjusting for sex, age, and CCI (Model 1) and for all variables in the table (Model 2)

When comparing hepatitis B individuals with and without NA-treatment, the risk for severe COVID-19 was increased in the NA-treated group in both the univariable (OR 1.954, 95% CI 1.473-2.560) and the multivariable analysis (OR 1.696 95% CI 1.218-2.334) (Table 4). In this analysis age, co-morbidities, lower income and education were associated with severe COVID-19, whereas COVID-19 vaccination had a protective effect 0.175 (0.123-0.243, p<0.001). Region of birth was not associated with more severe disease in the uni- or multivariable analysis 1.027 (0.743-1.456, p=0.878) vs 1.031 (0.707-1.537, p=0.878).

**Table 4.** Severe COVID-19 with or without NA-treatment. Logistic regression modelling the odds of severe COVID-19 in individuals with Hepatitis B (with and without N-analog treatment). Separate univariable models for each independent variable, and multivariable models adjusting for all variables in the table (Model 2)

| COVID-19 severity |  | Non-severe | Severe | OR (univariable) | OR (multivariable) |
| --- | --- | --- | --- | --- | --- |
| **NA-treatment** | No | 4918 (94.5) | 288 (5.5) | - | - |
|  | Yes | 603 (89.7) | 69 (10.3) | 1.954 (1.473-2.560, p<0.001) | 1.696 (1.218-2.334, p=0.001) |
| **Sex** | Male | 2769 (92.6) | 220 (7.4) | - | - |
|  | Female | 2752 (95.3) | 137 (4.7) | 0.627 (0.502-0.779, p<0.001) | 0.736 (0.573-0.941, p=0.015) |
| **Age (yrs)** | Mean (SD) | 43.8 (11.9) | 58.7 (13.0) | 1.095 (1.086-1.105, p<0.001) | 1.089 (1.078-1.100, p<0.001) |
| **wCCI_cat** | 0-2 | 4768 (95.7) | 213 (4.3) | - | - |
|  | 3-4 | 278 (85.5) | 47 (14.5) | 3.785 (2.672-5.264, p<0.001) | 1.301 (0.865-1.922, p=0.196) |
|  | 5+ | 475 (83.0) | 97 (17.0) | 4.571 (3.521-5.900, p<0.001) | 2.356 (1.723-3.200, p<0.001) |
| **COVID-19 vaccination** | No | 3288 (91.4) | 308 (8.6) | - | - |
|  | Yes | 2233 (97.9) | 49 (2.1) | 0.234 (0.171-0.315, p<0.001) | 0.175 (0.123-0.243, p<0.001) |
| **Education** | Primary Education | 1377 (91.2) | 133 (8.8) | - | - |
|  | Secondary Education | 1764 (94.0) | 112 (6.0) | 0.657 (0.506-0.853, p=0.002) | 0.861 (0.643-1.152, p=0.314) |
|  | Higher Education | 2116 (96.7) | 73 (3.3) | 0.357 (0.265-0.477, p<0.001) | 0.636 (0.457-0.880, p=0.007) |
|  | Missing or unknown | 264 (87.1) | 39 (12.9) | 1.529 (1.034-2.217, p=0.029) | 1.044 (0.623-1.710, p=0.868) |
| **Disp. Income** | Lowest | 2718 (91.9) | 240 (8.1) | - | - |
|  | Middle | 1869 (96.3) | 71 (3.7) | 0.430 (0.326-0.561, p<0.001) | 0.544 (0.400-0.732, p<0.001) |
|  | Highest | 912 (95.7) | 41 (4.3) | 0.509 (0.358-0.707, p<0.001) | 0.574 (0.390-0.826, p=0.004) |
|  | Missing or unknown | 22 (81.5) | 5 (18.5) | 2.574 (0.856-6.343, p=0.059) | 1.362 (0.346-4.709, p=0.640) |
| **Region of birth** | Northern Europe | 649 (94.1) | 41 (5.9) | - | - |
|  | Rest of the world | 4872 (93.9) | 316 (6.1) | 1.027 (0.743-1.456, p=0.878) | 1.031 (0.707-1.537, p=0.878) |

NA, nucleos(t)ide analogue; wCCI, weighted Charlson Comorbidity Index; N, number; OR, odds ratio; N.
Europe, yrs, years;

## Discussion

In this nationwide cohort of patients with CHB in Sweden, tenofovir use was associated with lower odds of severe COVID-19 compared with other NAs in unadjusted analyses. However, after adjustment for demographic, clinical, and socioeconomic factors, the association was attenuated and no longer statistically significant, suggesting that the apparent protective effect is uncertain and was largely explained by confounding from age, comorbidities, and vaccination status. We observed an increased risk of severe COVID-19 in CHB patients receiving NAs compared to those without treatment. This finding is not unexpected, as the NA-treated group consists of patients with more advanced liver disease and a higher burden of cirrhosis, which is a well-known risk for severe COVID-19 (28).

Our results align with recent evidence questioning the clinical relevance of tenofovir in attenuating COVID-19 severity. Observational studies early in the pandemic suggested that tenofovir, particularly TDF, might reduce the risk of severe COVID-19 in individuals with HIV or CHB(18, 21, 29). However, subsequent evidence has been inconsistent. For example, a large study from China found no significant difference in COVID-19 severity between CHB patients treated with tenofovir and those treated with entecavir (30). A smaller Spanish study reported a lower risk of severe disease among tenofovir-treated patients compared to those on entecavir, (17) and a randomised control pilot trial from 2024 reported lower risk for hospitalised patients treated with TAF, although the authors emphasized the need for larger, confirmatory studies(31). Such discrepancies may reflect differences in patient populations, sample size, and study design. As the largest study of its kind to date, our analysis provides greater statistical power and population-level precision. Importantly, the comprehensive Swedish registers enabled us to account for comorbidities and socioeconomic factors, which appear to drive much of the observed association.

In Sweden most hepatitis B patients are treated with tenofovir or entecavir according to national guidelines(16). From a mechanistic perspective, preclinical data have suggested that tenofovir can inhibit SARS-CoV-2 replication through interaction with the viral RNA-dependent RNA polymerase (RdRp), likely through the incorporation of its active triphosphate form into viral RNA, thereby impairing polymerase function (22, 32, 33) While both tenofovir and entecavir used to treat hepatitis B, are highly effective at suppressing HBV replication, only tenofovir has demonstrated potential RdRp inhibition *in vitro*, raising the question of whether its use might offer dual protection in HBV-infected individuals during the ongoing pandemic. Beyond direct antiviral effects, tenofovir may also influence host immune responses, including modulation of interferon-stimulated genes and innate immunity, which could theoretically contribute to improved outcomes. Entecavir does not share these properties, which has been suggested as a potential explanation for differences in clinical impact reported in some studies (22, 34).

However, our findings, together with other studies, (20, 25, 30, 35, 36) indicate that any such effects are not sufficient to translate into clinically meaningful protection against severe COVID-19. Instead, established risk factors including age, comorbidity burden, and vaccination status, remain the dominant determinants of severe COVID-19 outcomes in this group. Notably, our study underscores the critical role of vaccination in preventing severe COVID-19 among patients with chronic HBV. Vaccination was associated with a protective effect, significantly reducing the odds of severe disease. Despite this, vaccination coverage in our cohort was relatively low, with less than half of patients having received COVID-19 vaccination. Given the elevated baseline risk of severe outcomes in this population, improving vaccine coverage should therefore be a priority in the management of patients with CHB, especially in those with comorbidities.

In the adjusted analyses, we also observed an apparent protective association for individuals born outside Northern Europe. This finding should be interpreted with caution. Given the relatively small number of severe COVID-19 cases in this subgroup, even one or two individuals can substantially influence the estimates. Furthermore, previous large studies have not demonstrated a protective effect of migration background; on the contrary, immigrant populations have frequently been shown to experience higher risks of severe COVID-19 due to socioeconomic disadvantage, greater exposure, and healthcare barriers (37–39). Our finding therefore most likely reflects random variation rather than a true protective effect.

Strengths of our study include the nationwide design, large sample size, exclusion of patients with HIV, HCV, or HDV co-infections, and adjustment for age, sex, comorbidities, COVID-19 vaccination and socioeconomic covariates. Limitations include the observational design, potential residual confounding by treatment indication, and incomplete capture of liver disease stage. Furthermore, our study period encompassed different phases of the pandemic, including changing viral variants and vaccine rollout, which may have influenced outcomes.

In conclusion, this large national cohort study does not support a significant protective effect of tenofovir against severe COVID-19 in patients with CHB after adjustment for confounders. The consistently strong protective effect of vaccination, combined with the observed low vaccination coverage, highlights a need for targeted vaccination strategies in this group. Optimizing vaccine uptake and managing comorbidities remain the most effective strategies to reduce COVID-19 morbidity and mortality among patients living with CHB.

## Data Availability

No data was generated by this study. The following existing data sources were used: Healthcare registries from the Swedish National Board of Health and Welfare (Patient Registry, Cause of Death Registry), Swedish Public Health Agency, Statistics Sweden. Data is available from these registry stakeholder following ethical approval by the Swedish Ethical Review Board.

## Abbreviations

CHB: chronic hepatitis b
NA: nucleos(t)ide analogue
COVID-19: coronavirus disease 2019
HCV: hepatitis C virus
HDV: hepatitis delta virus
HBV: hepatitis B virus
OR: odds ratio
WHO: world health organisation
LAM: lamivudine,
ADV: adefovir dipivoxil
ETV: entecavir
TBV: telbivudine
TDF: tenofovir disoproxil fumarate
TAF: tenofovir alafenamide
IFNa: interferon alpha
PINs: personal identification numbers
LISA: Longitudinal Integrated Database for Health Insurance and Labor Market Studies
wCCI: weighted Charlson Comorbidity Index

## Supporting information

S1 Table. Countries divided in regions according to ISO3166

S2 Table. Baseline characteristics of non-severe and Severe COVID-19

